# Guillain-Barré Syndrome after COVID-19 Vaccination in the Vaccine Safety Datalink

**DOI:** 10.1101/2021.12.03.21266419

**Authors:** Kayla E. Hanson, Kristin Goddard, Ned Lewis, Bruce Fireman, Tanya R. Myers, Nandini Bakshi, Eric Weintraub, James G. Donahue, Jennifer C. Nelson, Stan Xu, Jason M. Glanz, Joshua T.B. Williams, Jonathan D. Alpern, Nicola P. Klein

## Abstract

**Importance:** Post-authorization monitoring of vaccines in a large population can detect rare adverse events not identified in clinical trials including Guillain-Barré syndrome (GBS). GBS has a background rate of 1-2 per 100,000 person-years.

**Objective:** To 1) describe cases and incidence of GBS following COVID-19 vaccination, and 2) assess the risk of GBS after vaccination for Ad.26.COV2.S (Janssen) and mRNA vaccines.

**Design:** Interim analysis of surveillance data from the Vaccine Safety Datalink.

**Setting:** Eight participating integrated healthcare systems in the United States.

**Participants:** 10,158,003 individuals aged ≥12 years.

**Exposures:** Receipt of Ad.26.COV2.S, BNT162b2 (Pfizer-BioNTech), or mRNA-1273 (Moderna) COVID-19 vaccine.

**Main Outcomes and Measures:** GBS with symptom onset in the 1-84 days after vaccination as confirmed by medical record review and adjudication. Descriptive characteristics of confirmed cases, GBS incidence rates during postvaccination risk intervals after each type of vaccine compared to the background rate, rate ratios (RRs) comparing GBS incidence in the 1-21 vs. 22-42 days postvaccination, and RRs directly comparing risk of GBS after Ad.26.COV2.S vs. mRNA vaccination, using Poisson regression adjusted for age, sex, race/ethnicity, site, and calendar day.

**Results:** From December 13, 2020 through November 13, 2021, 14,723,318 doses of COVID-19 vaccines were administered, including 467,126 Ad.26.COV2.S, 8,573,823 BNT162b2, and 5,682,369 mRNA-1273 doses. Eleven cases of GBS after Ad.26.COV2.S were confirmed. The unadjusted incidence rate of confirmed cases of GBS per 100,000 person-years in the 1-21 days after Ad.26.COV2.S was 34.6 (95% confidence interval [CI]: 15.8-65.7), significantly higher than the background rate, and the adjusted RR in the 1-21 vs. 22-42 days following Ad.26.COV2.S was 6.03 (95% CI: 0.79-147.79). Thirty-four cases of GBS after mRNA vaccines were confirmed. The unadjusted incidence rate of confirmed cases per 100,000 person-years in the 1-21 days after mRNA vaccines was 1.4 (95% CI: 0.7-2.5) and the adjusted RR in the 1-21 vs. 22-42 days following mRNA vaccines was 0.56 (95% CI: 0.21-1.48). In a head-to-head comparison of Ad.26.COV2.S vs. mRNA vaccines, the adjusted RR was 20.56 (95% CI: 6.94-64.66).

**Conclusions and Relevance:** In this interim analysis of surveillance data of COVID-19 vaccines, the incidence of GBS was elevated after Ad.26.COV2.S. Surveillance is ongoing.

## Introduction

Three vaccine products are available in the United States (US) to prevent coronavirus disease 2019 (COVID-19) including BNT162b2 (Pfizer-BioNTech), mRNA-1273 (Moderna), and Ad.26.COV2.S (Janssen).^1-3^ BNT162b2 and mRNA-1273 are both messenger RNA (mRNA) vaccines and administered as 2-dose primary series, whereas Ad.26.COV2.S is a replication-incompetent adenoviral vector vaccine and administered as a single dose primary series.

Guillain-Barré syndrome (GBS) is a rare neurological disorder with an incidence rate of 1-2 per 100,000 person-years.^4^ In July 2021, data from the Vaccine Adverse Event Reporting System (VAERS) indicated that the reporting rate of GBS was higher after Ad.26.COV2.S than after mRNA vaccines.^5^ On July 12, 2021, the Food and Drug Administration added a warning about GBS to the Ad.26.COV2.S vaccine fact sheet.

GBS is being monitored in the Vaccine Safety Datalink as part of ongoing rapid and prospective COVID-19 vaccine safety surveillance efforts.^6^ Our objectives for this report are to 1) describe GBS cases and incidence following COVID-19 vaccinations from December 13, 2020 through November 13, 2021, and 2) assess the risk of GBS after vaccination for Ad.26.COV2.S and mRNA vaccines.

## Methods

### Setting and Population

The Vaccine Safety Datalink is a collaboration between 9 US integrated healthcare systems and the Centers for Disease Control and Prevention (CDC).^7^ Eight data-contributing organizations (Kaiser Permanente: Colorado, Northern California, Northwest, Southern California, and Washington; Marshfield Clinic; HealthPartners; and Denver Health) have access to comprehensive medical records, including vaccinations, for 10,158,003 people aged ≥12 years as of November 10, 2021.

### Study Design

Since December 2020, the Vaccine Safety Datalink has been conducting safety surveillance of COVID-19 vaccines, monitoring GBS and 22 other pre-specified, serious outcomes after COVID-19 vaccines on a weekly basis. Methods are described in the study protocol (https://www.cdc.gov/vaccinesafety/ensuringsafety/monitoring/emergencypreparedness/index.html) and in a prior publication that includes interim results for mRNA vaccines.^6^ In brief, weekly analyses compared outcome incidence *observed* during a risk interval after vaccination (1-21 days or 1-42 days) with outcome incidence *expected*. The *expected* was derived from a) vaccinated comparators who were concurrently (on the same day) in a postvaccination comparison interval (22-42 days or 43-84 days, respectively) (“vaccinated concurrent comparators”), and in separate analyses from b) unvaccinated comparators who were concurrently unvaccinated (“unvaccinated concurrent comparators”). Individuals who received 2 doses of an mRNA vaccine contributed to these analyses when they were 1-21 days after dose 1 and again when they were 1-21 days after dose 2. However, receipt of dose 2 resulted in censoring of follow-up time after dose 1, therefore most comparison time in the vaccinated concurrent comparator analyses was after dose 2. This surveillance approach used 2 postvaccination risk intervals, 1-21 days and 1-42 days. The 1-21 day risk interval allowed for timelier analyses and avoids bias introduced from the short interval between doses for mRNA vaccines. However, a 1-42 day risk interval was also used, since this interval is often used in vaccine safety studies of GBS and other outcomes.^8^

By protocol, vaccinated concurrent comparator analyses were considered primary due to concerns about possible bias from unmeasured differences between vaccinated and unvaccinated individuals. However, the number of unvaccinated comparators is substantially larger than vaccinated comparators for Ad.26.COV2.S surveillance, because a large proportion of Ad.26.COV2.S was administered over a relatively short time period. Additionally, the power of Ad.26.COV2.S surveillance is limited by the relatively small number of doses administered. When analyses with vaccinated comparators are underpowered, more consideration is given to analyses with unvaccinated comparators, and to additional supplementary analyses. Here, we report on our most recent weekly analyses with vaccinated and unvaccinated comparators (a) and b) above), we compare the incidence of GBS postvaccination to the incidence expected from prior studies, we evaluate the temporal clustering of GBS cases by the number of days from vaccination to the onset of GBS symptoms, and we perform head-to-head comparisons of GBS incidence after Ad.26.COV2.S with GBS incidence after mRNA vaccination. This activity was approved by the Institutional Review Boards (IRBs) with a waiver of informed consent at Kaiser Permanente: Colorado, Northern California, Northwest, Southern California, and Washington; Marshfield Clinic; HealthPartners; and Denver Health. CDC determined that this activity was public health surveillance (45 C.F.R. part 46.102(l)(2)) and thus did not require IRB review.

Potential cases of GBS were identified using International Classification of Diseases 10^th^ Revision (ICD-10) code G61.0 in the emergency department or inpatient setting, specifically when G61.0 first appeared in an individual’s record in the 1-84 days after any COVID-19 vaccine. Because disease onset may begin before a diagnosis is recorded in the medical record, potential cases with the ICD-10 code in the 85-98 days after vaccination were also reviewed. After review, all cases underwent adjudication according to the Brighton Collaboration criteria.^9^ Briefly, GBS cases meeting Brighton level 1 had the highest level of diagnostic certainty, while level 4 included cases of suspected GBS (i.e., insufficient information was available in the medical record to meet Brighton level 1-3). In this analysis, we considered Brighton level 1-4 cases as confirmed and sensitivity analyses were conducted excluding level 4 cases. Cases of GBS among unvaccinated concurrent comparators were identified using the same approach, but did not undergo medical record review and adjudication.

### Statistical Analysis

As previously described,^6^ primary weekly monitoring of GBS after each type of COVID-19 vaccine used conditional Poisson regression to compare individuals during a postvaccination risk interval with similar individuals who had been vaccinated earlier and were concurrently in a comparison interval. These analyses estimated rate ratios (i.e., ratios of incidence rates in a risk interval divided by incidence rates in a comparison interval), that were adjusted for 5-year age group, sex, race/ethnicity, site, and calendar day by conditioning the Poisson regression on strata (risk sets) defined by these factors. Similarly, conditional Poisson regression was used in supplementary weekly analyses to compare individuals in a postvaccination risk interval with similar individuals who were concurrently unvaccinated. 1-sided sequential testing was conducted weekly for vaccinated concurrent comparator analyses, with a signaling threshold of *P*<.0048 (pre-specified to keep the overall chance of making a type I error below .05 during 2 years of weekly analyses).

Although our primary weekly analyses of GBS have not met the criterion for a safety signal, concerns about GBS after Ad.26.COV2.S arose from supplementary analyses with unvaccinated comparators and from reports to VAERS. To further investigate we conducted additional analyses. We computed unadjusted incidence rates and 95% confidence intervals (CIs) of confirmed GBS in the 1-21 days and 1-42 days following COVID-19 vaccination per 100,000 person-years (PY) by vaccine type and compared them to the historical background rate of GBS (2 per 100,000 PY).^4^ We conducted head-to-head comparisons of the 21-day and 42-day risk intervals after Ad.26.COV2.S vs. the 21-day and 42-day risk intervals after mRNA vaccination, using conditional Poisson regression to estimate rate ratios (i.e., the ratio of GBS incidence after Ad.26.COV2.S divided by GBS incidence after mRNA vaccination), adjusted for age group, sex, race/ethnicity, site, and calendar day as described above. We also examined the temporal clustering of the GBS cases within 56 days of Ad.26.COV2.S by day of onset, using a scan statistic.^10^ All analyses of mRNA vaccines combined BNT162b2 and mRNA-1273, all analyses were 2-sided with a .05 level of significance unless otherwise specified, and all analyses were conducted using SAS version 9.4 (Cary, NC). This report includes COVID-19 vaccinations and GBS diagnoses through November 13, 2021.

## Results

### COVID-19 Vaccinations

From December 13, 2020 through November 13, 2021, 14,723,318 COVID-19 vaccines were administered to individuals ≥12 years of age including 467,126 first doses of Ad.26.COV2.S, 8,573,823 first and second doses of BNT162b2, and 5,682,369 first and second doses of mRNA-1273; mRNA vaccines accounted for 96.8% of all doses. Compared with mRNA vaccinees, a higher proportion of Ad.26.COV2.S vaccinees were male (45.9% vs. 53.9%) and a smaller proportion were ≥65 years of age (21.7% vs. 11.7%)(**Table 1**).

**Table 1.** COVID-19 Vaccine Doses According to Characteristics of the Vaccinee^a^.

| **Characteristic<sup>b</sup>** | **Ad.26.COV2.S**<br>**(N=467,126)** | **mRNA Vaccine<sup>c</sup>**<br>**(N=14,256,192)** |
| --- | --- | --- |
| Dose number |  |  |
| 1 | 467,126 (100.0%) | 7,243,740 (50.8%) |
| 2 | N/A | 7,012,452 (49.2%) |
| Sex |  |  |
| Male | 251,549 (53.9%) | 6,543,571 (45.9%) |
| Female | 215,577 (46.2%) | 7,712,621 (54.1%) |
| Age group |  |  |
| 12-17 years | N/A | 1,102,346 (7.7%) |
| 18-49 years | 250,526 (53.6%) | 6,656,291 (46.7%) |
| 50-64 years | 161,886 (34.7%) | 3,405,541 (23.9%) |
| 65-74 years | 36,719 (7.9%) | 1,885,175 (13.2%) |
| ≥75 years | 17,995 (3.9%) | 1,206,839 (8.5%) |
| Race/ethnicity<sup>d</sup> |  |  |
| White, non-Hispanic | 213,660 (45.7%) | 5,872,570 (41.2%) |
| Black, non-Hispanic | 31,043 (6.7%) | 861,612 (6.0%) |
| Asian, non-Hispanic | 54,253 (11.6%) | 2,105,692 (14.8%) |
| Native Hawaiian/Pacific<br>Islander, non-Hispanic | 2,722 (0.6%) | 90,325 (0.6%) |
| American Indian/Alaska<br>Native, non-Hispanic | 1,558 (0.3%) | 42,783 (0.3%) |
| Multiple/Other, non-Hispanic | 14,442 (3.1%) | 496,932 (3.5%) |
| Hispanic/Latino | 97,332 (20.8%) | 3,453,794 (24.2%) |
| Unknown | 52,116 (11.2%) | 1,332,484 (9.4%) |
| History of COVID-19 | 41,265 (8.8%) | 1,059,225 (7.4%) |

Abbreviations: N/A=not applicable.
<sup>a</sup>Unit of analysis is vaccine dose.
<sup>b</sup>Data are presented as number (percentage); percentages may not add to 100% due to rounding.
<sup>c</sup>Includes 8,573,823 doses of BNT162b2 and 5,682,369 doses of mRNA-1273.
<sup>d</sup>8 combined race/ethnicity categories were created from available race and ethnicity information. Persons were classified as Hispanic/Latino if categorized as having Hispanic/Latino ethnicity, regardless of whether or not race was known. Persons were classified as having unknown race/ethnicity if both race and ethnicity were missing.

### GBS after Ad.26.COV2.S

During the 1-84 days after Ad.26.COV2.S, 21 potential cases of GBS were identified: all were reviewed and adjudicated, and 11 (52%) were confirmed. Symptom onset ranged from 1 to 77 days after Ad.26.COV2.S with onset within 14 days of vaccination for 9 of 11 (82%) confirmed cases (**Figure 1A**). Scan statistics identified days 1-14 after vaccination as a statistically significant cluster (*P*=.003). Most patients were hospitalized (91%) and all were treated with intravenous immune globulin. Patients with confirmed GBS after Ad.26.COV2.S had a mean age of 50 years (range 32 to 63 years), most were male (82%) and non-Hispanic white (91%), and most had facial weakness or paralysis (91%) in addition to bilateral weakness or paralysis of the limbs (**Table 2**).

**Figure 1.**
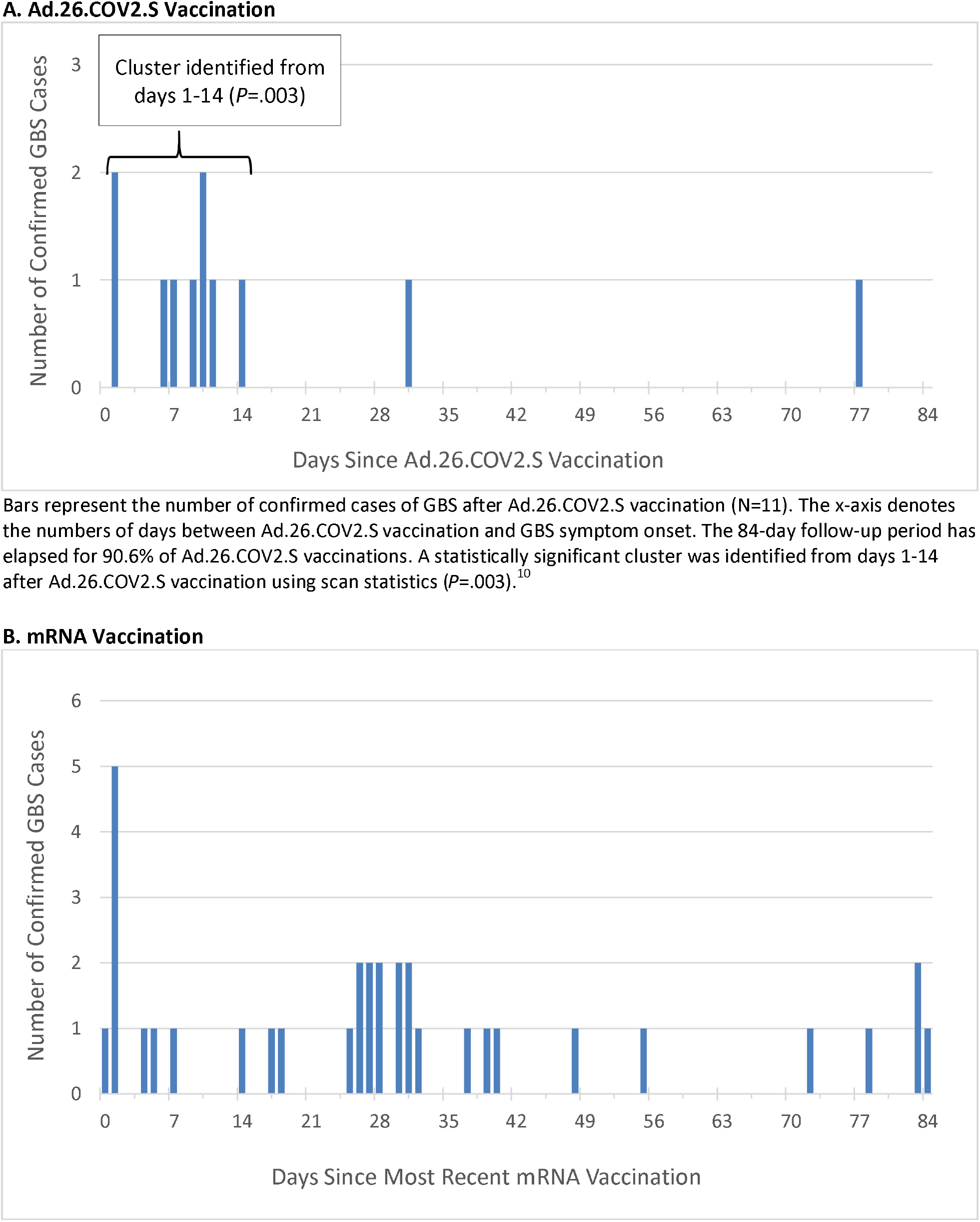
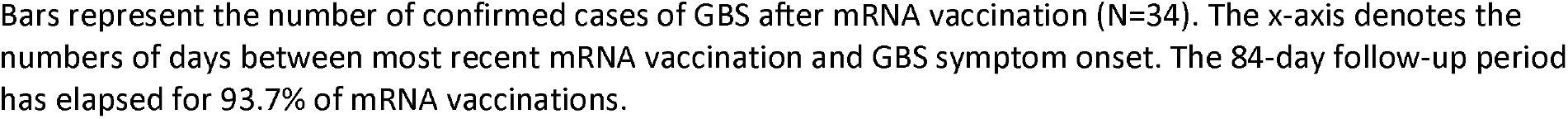
Timing of GBS Symptom Onset after COVID-19 Vaccination.

**Table 2.**
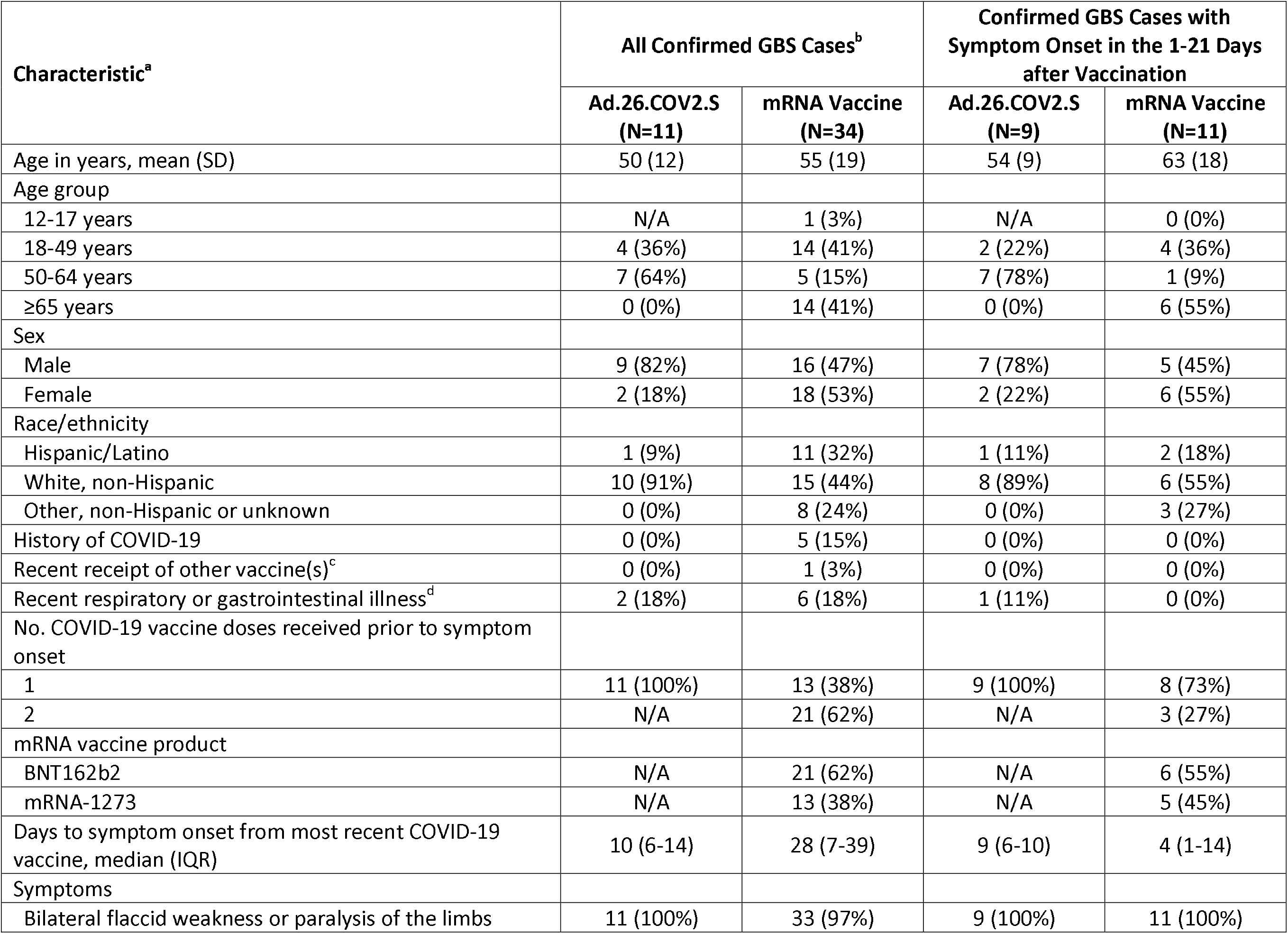
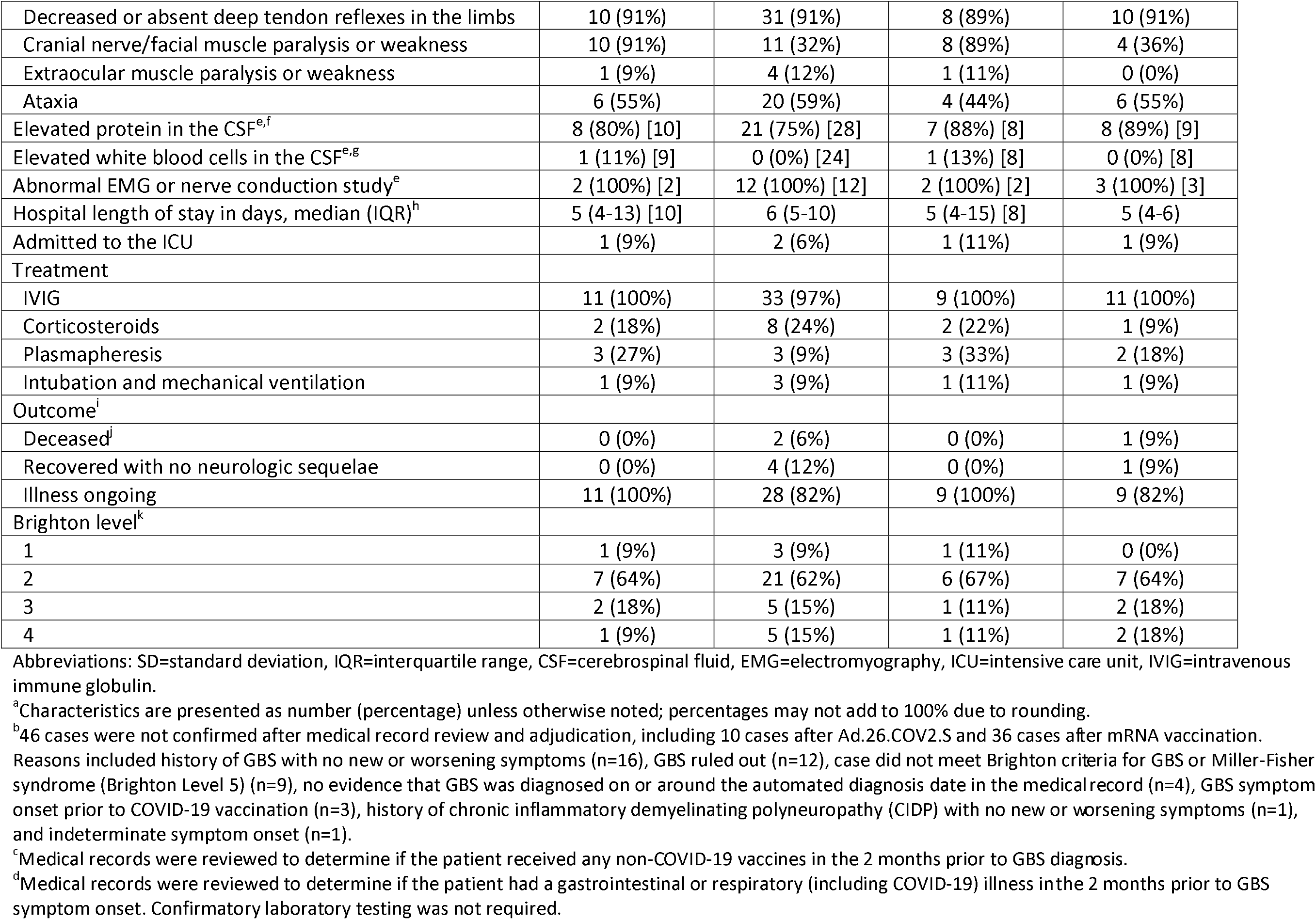
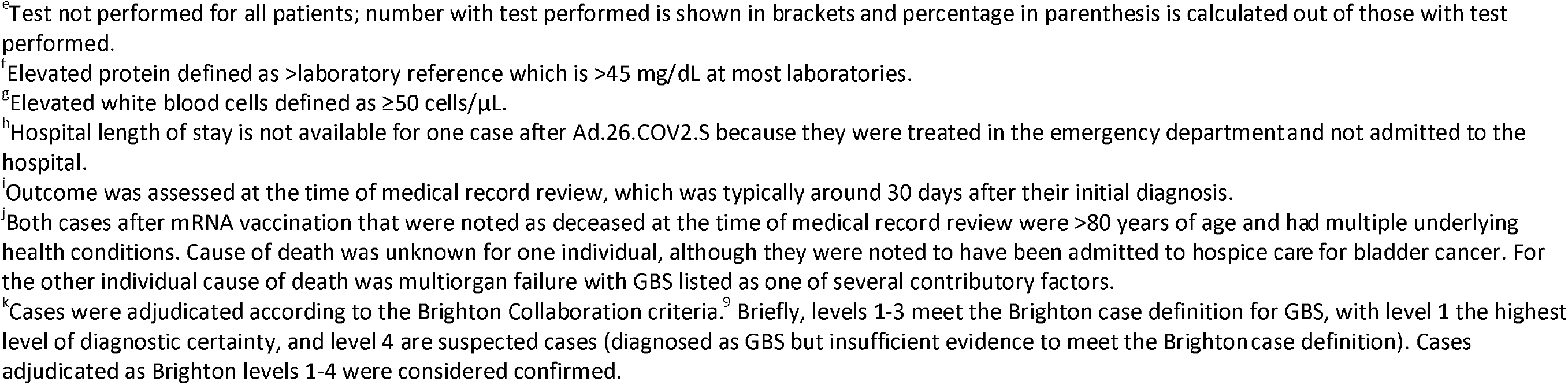
Characteristics of Confirmed GBS Cases after COVID-19 Vaccination.

| Characteristic<sup>a</sup> | All Confirmed GBS Cases<sup>b</sup> |  | Confirmed GBS Cases with Symptom Onset in the 1-21 Days after Vaccination |  |
| --- | --- | --- | --- | --- |
|  | Ad.26.COV2.S (N=11) | mRNA Vaccine (N=34) | Ad.26.COV2.S (N=9) | mRNA Vaccine (N=11) |
| Age in years, mean (SD) | 50 (12) | 55 (19) | 54 (9) | 63 (18) |
| Age group |  |  |  |  |
| 12-17 years | N/A | 1 (3%) | N/A | 0 (0%) |
| 18-49 years | 4 (36%) | 14 (41%) | 2 (22%) | 4 (36%) |
| 50-64 years | 7 (64%) | 5 (15%) | 7 (78%) | 1 (9%) |
| ≥65 years | 0 (0%) | 14 (41%) | 0 (0%) | 6 (55%) |
| Sex |  |  |  |  |
| Male | 9 (82%) | 16 (47%) | 7 (78%) | 5 (45%) |
| Female | 2 (18%) | 18 (53%) | 2 (22%) | 6 (55%) |
| Race/ethnicity |  |  |  |  |
| Hispanic/Latino | 1 (9%) | 11 (32%) | 1 (11%) | 2 (18%) |
| White, non-Hispanic | 10 (91%) | 15 (44%) | 8 (89%) | 6 (55%) |
| Other, non-Hispanic or unknown | 0 (0%) | 8 (24%) | 0 (0%) | 3 (27%) |
| History of COVID-19 | 0 (0%) | 5 (15%) | 0 (0%) | 0 (0%) |
| Recent receipt of other vaccine(s)<sup>c</sup> | 0 (0%) | 1 (3%) | 0 (0%) | 0 (0%) |
| Recent respiratory or gastrointestinal illness<sup>d</sup> | 2 (18%) | 6 (18%) | 1 (11%) | 0 (0%) |
| No. COVID-19 vaccine doses received prior to symptom onset |  |  |  |  |
| 1 | 11 (100%) | 13 (38%) | 9 (100%) | 8 (73%) |
| 2 | N/A | 21 (62%) | N/A | 3 (27%) |
| mRNA vaccine product |  |  |  |  |
| BNT162b2 | N/A | 21 (62%) | N/A | 6 (55%) |
| mRNA-1273 | N/A | 13 (38%) | N/A | 5 (45%) |
| Days to symptom onset from most recent COVID-19 vaccine, median (IQR) | 10 (6-14) | 28 (7-39) | 9 (6-10) | 4 (1-14) |
| Symptoms |  |  |  |  |
| Bilateral flaccid weakness or paralysis of the limbs | 11 (100%) | 33 (97%) | 9 (100%) | 11 (100%) |
| Decreased or absent deep tendon reflexes in the limbs | 10 (91%) | 31 (91%) | 8 (89%) | 10 (91%) |
| Cranial nerve/facial muscle paralysis or weakness | 10 (91%) | 11 (32%) | 8 (89%) | 4 (36%) |
| Extraocular muscle paralysis or weakness | 1 (9%) | 4 (12%) | 1 (11%) | 0 (0%) |
| Ataxia | 6 (55%) | 20 (59%) | 4 (44%) | 6 (55%) |
| Elevated protein in the CSF<sup>e,f</sup> | 8 (80%) [10] | 21 (75%) [28] | 7 (88%) [8] | 8 (89%) [9] |
| Elevated white blood cells in the CSF<sup>e,g</sup> | 1 (11%) [9] | 0 (0%) [24] | 1 (13%) [8] | 0 (0%) [8] |
| Abnormal EMG or nerve conduction study<sup>e</sup> | 2 (100%) [2] | 12 (100%) [12] | 2 (100%) [2] | 3 (100%) [3] |
| Hospital length of stay in days, median (IQR)<sup>h</sup> | 5 (4-13) [10] | 6 (5-10) | 5 (4-15) [8] | 5 (4-6) |
| Admitted to the ICU | 1 (9%) | 2 (6%) | 1 (11%) | 1 (9%) |
| Treatment |  |  |  |  |
| IVIG | 11 (100%) | 33 (97%) | 9 (100%) | 11 (100%) |
| Corticosteroids | 2 (18%) | 8 (24%) | 2 (22%) | 1 (9%) |
| Plasmapheresis | 3 (27%) | 3 (9%) | 3 (33%) | 2 (18%) |
| Intubation and mechanical ventilation | 1 (9%) | 3 (9%) | 1 (11%) | 1 (9%) |
| Outcome<sup>i</sup> |  |  |  |  |
| Deceased<sup>j</sup> | 0 (0%) | 2 (6%) | 0 (0%) | 1 (9%) |
| Recovered with no neurologic sequelae | 0 (0%) | 4 (12%) | 0 (0%) | 1 (9%) |
| Illness ongoing | 11 (100%) | 28 (82%) | 9 (100%) | 9 (82%) |
| Brighton level<sup>k</sup> |  |  |  |  |
| 1 | 1 (9%) | 3 (9%) | 1 (11%) | 0 (0%) |
| 2 | 7 (64%) | 21 (62%) | 6 (67%) | 7 (64%) |
| 3 | 2 (18%) | 5 (15%) | 1 (11%) | 2 (18%) |
| 4 | 1 (9%) | 5 (15%) | 1 (11%) | 2 (18%) |

Abbreviations: SD=standard deviation, IQR=interquartile range, CSF=cerebrospinal fluid, EMG=electromyography, ICU=intensive care unit, IVIG=intravenous immune globulin.
<sup>a</sup>Characteristics are presented as number (percentage) unless otherwise noted; percentages may not add to 100% due to rounding.
<sup>b</sup>46 cases were not confirmed after medical record review and adjudication, including 10 cases after Ad.26.COVS.2 and 36 cases after mRNA vaccination.
Reasons included history of GBS with no new or worsening symptoms (n=16), GBS ruled out (n=12), case did not meet Brighton criteria for GBS or Miller-Fisher syndrome (Brighton Level 5) (n=9), no evidence that GBS was diagnosed on or around the automated diagnosis date in the medical record (n=4), GBS symptom onset prior to COVID-19 vaccination (n=3), history of chronic inflammatory demyelinating polyneuropathy (CIDP) with no new or worsening symptoms (n=1), and indeterminate symptom onset (n=1).
<sup>c</sup>Medical records were reviewed to determine if the patient received any non-COVID-19 vaccines in the 2 months prior to GBS diagnosis.
<sup>d</sup>Medical records were reviewed to determine if the patient had a gastrointestinal or respiratory (including COVID-19) illness in the 2 months prior to GBS symptom onset. Confirmatory laboratory testing was not required.
<sup>e</sup>Test not performed for all patients; number with test performed is shown in brackets and percentage in parenthesis is calculated out of those with test performed.
<sup>f</sup>Elevated protein defined as >laboratory reference which is >45 mg/dL at most laboratories.
<sup>g</sup>Elevated white blood cells defined as $\geq 50$ cells/ $\mu$ L.
<sup>h</sup>Hospital length of stay is not available for one case after Ad.26.COV2.S because they were treated in the emergency department and not admitted to the hospital.
<sup>i</sup>Outcome was assessed at the time of medical record review, which was typically around 30 days after their initial diagnosis.
<sup>j</sup>Both cases after mRNA vaccination that were noted as deceased at the time of medical record review were >80 years of age and had multiple underlying health conditions. Cause of death was unknown for one individual, although they were noted to have been admitted to hospice care for bladder cancer. For the other individual cause of death was multiorgan failure with GBS listed as one of several contributory factors.

The unadjusted incidence rate of confirmed cases of GBS per 100,000 PY was 34.6 (95% CI: 15.8-65.7) during the 1-21 days after Ad.26.COV2.S and 19.2 (95% CI: 9.2-35.4) during the 1-42 days after Ad.26.COV2.S; both estimates were significantly higher than the background rate of GBS (*P*<.001) (**Table 3**). When excluding Brighton level 4 cases, incidence rates were lower (30.8 and 17.3 per 100,000 PY, respectively) but still significantly higher than the background rate (*P*<.001).

**Table 3.** Incidence Rate of Confirmed GBS in the 1-21 Days and 1-42 Days after COVID-19 Vaccination.

| Vaccine Type | Risk Window | Including BL 4 Cases<sup>a</sup> | No. GBS Cases | No. Vaccine Doses<sup>b</sup> | No. Person-Years<sup>c</sup> | Unadjusted Incidence Rate per Million Doses (95% CI) | Unadjusted Incidence Rate per 100,000 PY (95% CI) | 2-sided P value<sup>d</sup> |
| --- | --- | --- | --- | --- | --- | --- | --- | --- |
| Ad.26.COV2.S | 1-21 days | Yes | 9 | 452,105 | 25,994 | 19.9 (9.1-37.8) | 34.6 (15.8-65.7) | <.001 |
|  |  | No | 8 | 452,105 | 25,994 | 17.7 (7.6-34.9) | 30.8 (13.3-60.6) | <.001 |
|  | 1-42 days | Yes | 10 | 452,105 | 51,987 | 22.1 (10.6-40.7) | 19.2 (9.2-35.4) | <.001 |
|  |  | No | 9 | 452,105 | 51,987 | 19.9 (9.1-37.8) | 17.3 (7.9-32.9) | <.001 |
| mRNA Vaccine | 1-21 days | Yes | 11 | 13,952,901 | 792,869 | 0.8 (0.4-1.4) | 1.4 (0.7-2.5) | .27 |
|  |  | No | 9 | 13,952,901 | 792,869 | 0.6 (0.3-1.2) | 1.1 (0.5-2.2) | .09 |
|  | 1-42 days | Yes | 26 | 13,952,901 | 1,264,707 | 1.9 (1.2-2.7) | 2.1 (1.3-3.0) | .94 |
|  |  | No | 23 | 13,952,901 | 1,264,707 | 1.6 (1.0-2.5) | 1.8 (1.2-2.7) | .74 |

Abbreviations: BL=Brighton level, No.=number, CI=confidence interval, PY=person-years.
<sup>a</sup>Sensitivity analyses were conducted excluding Brighton level 4 cases (suspected cases).
<sup>b</sup>Includes vaccine doses administered through October 2, 2021 to allow for 42 days of follow-up.
<sup>c</sup>Follow-up time after dose 1 of either mRNA vaccine was censored after receipt of dose 2.
<sup>d</sup>The background rate of GBS is 1-2 per 100,000 person-years.<sup>4</sup> Exact Poisson regression was used to compare the observed number of GBS cases with the expected number of cases, which was derived from a background rate of 2 per 100,000 person-years and the observed number of person-years.

In weekly surveillance using vaccinated concurrent comparators, as of November 16, 2021, the RR of confirmed GBS, adjusted for age, sex, race/ethnicity, site, and calendar day in the 1-21 vs. 22-42 days following Ad.26.COV2.S was 6.03 (95% CI: 0.79-147.79, 2-sided *P*=.09, 1-sided *P*=.08). The adjusted RR of confirmed GBS in the 1-42 vs. 43-84 days following Ad.26.COV2.S was 8.64 (95% CI: 1.18-207.32, 2-sided *P*=.03, 1-sided *P*=.03). Neither result met the prespecified signaling criteria of a 1-sided *P*<.0048.

In supplemental weekly analyses with unvaccinated concurrent comparators, the adjusted RR of GBS was 10.57 (95% CI: 5.15-20.16, *P*<.001) in the 1-21 days following Ad.26.COV2.S and 10.05 (95% CI: 5.75-16.96, *P*<.001) in the 1-42 days following Ad.26.COV2.S.

### GBS after mRNA Vaccines

During the 1-84 days after first and second doses of mRNA vaccines, 71 potential cases of GBS were identified: 70 (99%) were reviewed and adjudicated (1 pending), and 34 (49%) were confirmed, including 21 after BNT162b2 and 13 after mRNA-1273. Patients with confirmed GBS after mRNA vaccines had a mean age of 55 years (range 15 to 92 years) (**Table 2**) and symptom onset ranged from 0 to 84 days after vaccination (**Figure 1B**). Eleven cases (32%) had symptom onset in the 1-21 days after vaccination and 15 cases (44%) had symptom onset in the 22-42 days after vaccination.

The unadjusted incidence rate of confirmed cases of GBS per 100,000 PY in the 21 days after mRNA vaccines was 1.4 (95% CI: 0.7-2.5) and did not differ from the background rate of GBS (*P*=.27) (**Table 3**). Results were similar when excluding Brighton level 4 cases and when using a 42-day risk window, with incidence rates ranging from 1.1 to 2.1.

In weekly surveillance conducted on November 16, 2021, the adjusted RR of confirmed GBS in the 1-21 vs. 22-42 days following mRNA vaccination was 0.56 (95% CI: 0.21-1.48, 2-sided *P*=.25, 1-sided *P*=.93).

In supplemental weekly analyses with unvaccinated concurrent comparators, the adjusted RR of GBS was 0.83 (95% CI: 0.50-1.33, *P*=.45) in the 1-21 days following mRNA vaccination and 0.85 (95% CI: 0.57-1.27, *P*=.44) in the 1-42 days following mRNA vaccination.

### Head-to-Head Comparisons

During the 1-21 days after vaccination, the adjusted RR of confirmed GBS after Ad.26.COV2.S vs. mRNA vaccination was 20.56 (95% CI: 6.94-64.66, *P*<.001), with 15.5 excess GBS cases in the risk interval per million Ad.26.COV2.S vaccinees (**Table 4**). When using a 1-42 day risk interval the adjusted RR was lower, 11.46, otherwise results were similar.

**Table 4.** Head-to-Head Comparisons of Confirmed GBS Incidence after Ad.26.COV2.S vs. mRNA Vaccination.

| **Risk Window** | **No. GBS Cases after Ad.26.COV2.S** | **No. GBS Cases after mRNA Vaccination<sup>a</sup>** | **Adjusted RR (95% CI)<sup>b</sup>** | **2-sided *P* value** | **Excess Cases in Risk Interval per Million Doses** |
| --- | --- | --- | --- | --- | --- |
| 1-21 days | 9 | 8 | 20.56 (6.94-64.66) | <.001 | 15.5 |
| 1-42 days | 10 | 21 | 11.46 (4.83-26.16) | <.001 | 17.5 |

Abbreviations: No.=number, RR=rate ratio, CI=confidence interval.
<sup>a</sup>Not all confirmed cases of GBS after mRNA vaccination are included in this analysis, such as cases that occurred prior to the authorization of Ad.26.COV2.S.
<sup>b</sup>Adjusted for 5-year age group, sex, race/ethnicity, site, and calendar day.

## Discussion

In interim analyses conducted in a large population-based surveillance system that includes medical record review of all potential GBS cases after COVID-19 vaccination, findings were consistent with an increased risk of GBS after Ad.26.COV2.S. The incidence of GBS in the 21 days after Ad.26.COV2.S was 34.6 per 100,000 person-years, which was substantially greater than the expected background rate of 1-2 per 100,000 person-years.^4^ GBS incidence in the 21 days after mRNA vaccination was 1.4 per 100,000 person-years, similar to the overall expected background rate. In an adjusted head-to-head comparison, GBS incidence during the 21 days after Ad.26.COV2.S was 20.6 times higher than the GBS incidence during the 21 days after mRNA vaccination, amounting to 15.5 excess cases per million Ad.26.COV2.S vaccinees. The majority of cases of GBS after Ad.26.COV2.S occurred during the 1-21 day risk interval, with the period of most increased risk in the 1-14 days after Ad.26.COV2.S.

None of the primary analyses conducted in routine weekly safety surveillance of GBS after Ad.26.COV2.S met the criterion for a safety signal, however these analyses have low power not only because the uptake of Ad.26.COV2.S has been low, but also because Ad.26.COV2.S uptake was concentrated in a brief calendar period so that relatively few vaccinated concurrent comparators were available while vaccinees were in their risk interval. The rate ratio estimate of 6.0 in the most recent primary weekly analyses of the 21-day risk interval after Ad.26.COV2.S is surrounded by a wide confidence interval extending from <1.0 to >100. Because our supplementary analyses are much more powerful, they should be given more consideration, consistent with the study’s protocol, but they should be interpreted with caution because of concerns about unmeasured differences between vaccinated and unvaccinated individuals.

Nevertheless, our finding of an elevated risk of GBS after Ad.26.COV2.S is consistent with a recent observed-to-expected analysis in VAERS which found that the GBS reporting rate after Ad.26.COV2.S exceeded the background rate.^11^ Our analyses also include GBS cases confirmed by medical record review and adjudication, and thus provide validation of the preliminary VAERS findings based on presumptive GBS.

Interestingly, nearly all patients with GBS after Ad.26.COV2.S had facial weakness or paralysis. Although GBS typically presents with weakness in the limbs, facial weakness occurs in 20-70% of cases.^12,13^ Reports from the United Kingdom and India describing cases of GBS following ChAdOx1 (AstraZeneca) COVID-19 vaccine, another adenoviral vector vaccine, have also observed facial weakness.^14,15^

In contrast to Ad.26.COV2.S, the unadjusted incidence rates of GBS in the 1-21 days and 1-42 days after mRNA vaccines were similar to the published background rate.^4^ In weekly analyses, the incidence of GBS following mRNA vaccination was also not significantly higher in the 1-21 days postvaccination compared with 22-42 days postvaccination, a finding consistent with similar results reported through June 22, 2021 from this same surveillance.^6^ These updated results provide further evidence that mRNA vaccines do not appear to be associated with GBS.

### Limitations

This study had several limitations. First, as mentioned above, substantially fewer doses of Ad.26.COV2.S were administered relative to mRNA vaccines, resulting in reduced statistical power and wide confidence intervals for some analyses. Second, in this observational study recipients of Ad.26.COV2.S may have differed from recipients of mRNA vaccines in unknown ways that affect GBS risk but were not adjusted for in analyses. Third, the incidence rate of confirmed GBS during the COVID-19 pandemic has not been established, and may differ from pre-pandemic background rates. Fourth, we could not identify subgroups who may be at greatest risk for GBS after Ad.26.COV2.S given the small number of confirmed GBS cases identified in this surveillance. Fifth, this analysis only included GBS cases after primary COVID-19 vaccination and results may not be generalizable to additional or booster doses.

## Conclusions

In this interim analysis of surveillance data of COVID-19 vaccines, findings were consistent with an elevated risk of GBS after primary Ad.26.COV2.S vaccination. Surveillance is ongoing.

## Data Availability

VSD data are not publicly available but access may be granted through collaboration with VSD sites or the VSD data sharing program. Please visit https://www.cdc.gov/vaccinesafety/ensuringsafety/monitoring/vsd/accessing-data.html for more information.

## Author Contributions

Ms. Hanson had full access to all of the data in the study and takes responsibility for the integrity of the data and the accuracy of the data analysis.

*Concept and design:* Hanson, Klein, Goddard, Lewis, Fireman, Weintraub, Xu, Williams, Nelson, Glanz

*Acquisition, analysis, or interpretation of data:* Hanson, Donahue, Klein, Goddard, Bakshi, Lewis, Fireman, Weintraub, Myers, Xu, Williams, Nelson, Glanz, Alpern

*Drafting of the manuscript:* Hanson, Klein, Goddard

*Critical revision of the manuscript for important intellectual content:* Donahue, Klein, Goddard, Bakshi, Lewis, Fireman, Weintraub, Myers, Xu, Williams, Nelson, Glanz, Alpern

*Statistical analysis:* Hanson, Lewis

*Obtained funding:* Hanson, Donahue, Klein, Goddard

*Administrative, technical, or material support:* Hanson, Donahue, Klein, Goddard, Lewis, Fireman, Weintraub, Xu, Williams, Nelson, Glanz, Alpern

## Disclaimers

The findings and conclusions in this paper are those of the authors and do not necessarily represent the official position of the CDC. Mention of a product or company name is for identification purposes only and does not constitute endorsement by CDC.

## Acknowledgments

We thank Ousseny Zerbo, PhD (Kaiser Permanente Vaccine Study Center, Kaiser Permanente Northern California) for contributions to study design and concurrent comparator analyses. We thank Burney Kieke, MA (Marshfield Clinic Research Institute) for contributions to observed-to-expected analyses. We thank Malini DeSilva, MD (HealthPartners Institute), Elyse Kharbanda, MD, MPH (HealthPartners Institute) and Allison Naleway, PhD (Center for Health Research, Kaiser Permanente Northwest) for oversight of data collection and interpretation of data. We thank Rachael Burganowski, MS (Kaiser Permanente Washington Health Research Institute), Bradley Crane, MS (Center for Health Research, Kaiser Permanente Northwest), Sungching Glenn, MS (Research and Evaluation, Kaiser Permanente Southern California), Tat’Yana Kenigsberg, MPH (Immunization Safety Office, Centers for Disease Control and Prevention), Yingbo Lou, MS (Ambulatory Care Services, Denver Health), John Mayer, PhD (Marshfield Clinic Research Institute), Erica Scotty, MS (Marshfield Clinic Research Institute), Gabriela Vazquez Benitez, PhD (HealthPartners Institute), Arnold Yee, BS (Kaiser Permanente Vaccine Study Center, Kaiser Permanente Northern California), and Jingyi Zhu, PhD (HealthPartners Institute), for their contributions to data collection and preparation. We thank Dawn Asamura, BS (Research and Evaluation, Kaiser Permanente Southern California), Radha Bathala, MS (Research and Evaluation, Kaiser Permanente Southern California), Nancy Canul-Jauriga (Research and Evaluation, Kaiser Permanente Southern California), Alexander Carruth (Research and Evaluation, Kaiser Permanente Southern California), Jennifer Covey, BS (Kaiser Permanente Washington Health Research Institute), Susie Flores, RN (Research and Evaluation, Kaiser Permanente Southern California), Joy Gelfond (Research and Evaluation, Kaiser Permanente Southern California), Stacy Harsh, BSN, RN (Center for Health Research, Kaiser Permanente Northwest), Linda Heeren, BS (Marshfield Clinic Research Institute), Sunhea Kim, MPH (Research and Evaluation, Kaiser Permanente Southern California), Kate Kurlandsky, BA (Center for Health Systems Research, Denver Health), Jose Pio, MD, MPH (Research and Evaluation, Kaiser Permanente Southern California), Pat Ross, BA (Kaiser Permanente Vaccine Study Center, Kaiser Permanente Northern California), Marycania Saparudin, MPH (Research and Evaluation, Kaiser Permanente Southern California), Karen Schenk, BA (Research and Evaluation, Kaiser Permanente Southern California), Sarah Simmons, MPH (Research and Evaluation, Kaiser Permanente Southern California), Laura Sirikulvadhana, MPH (Research and Evaluation, Kaiser Permanente Southern California), and Melena Taylor, BA (Research and Evaluation, Kaiser Permanente Southern California), for their contributions to medical record review. We also thank Laurie Aukes, RN (Kaiser Permanente Vaccine Study Center, Kaiser Permanente Northern California), Jonathan Block, MEd (Ambulatory Care Services, Denver Health), Cheryl Carlson, MPH (Research and Evaluation, Kaiser Permanente Southern California), Stephanie Irving, MHS (Center for Health Research, Kaiser Permanente Northwest), Mara Kalter, MA (Center for Health Research, Kaiser Permanente Northwest), Tia Kauffman, MPH (Center for Health Research, Kaiser Permanente Northwest), Erika Kiniry, MPH (Kaiser Permanente Washington Health Research Institute), Leslie Kuckler, MPH (HealthPartners Institute), Denison Ryan, MPH (Research and Evaluation, Kaiser Permanente Southern California), and Lina Sy, MPH (Research and Evaluation, Kaiser Permanente Southern California), for their contributions to overall project management. All non-CDC personnel received financial compensation through CDC Vaccine Safety Datalink grant funding for their work on this project. CDC personnel were not compensated for their role in the study.

